# Mathematical Modeling and a Month Ahead Forecast of the Coronavirus Disease 2019 (COVID-19) Pandemic: An Indian Scenario

**DOI:** 10.1101/2020.09.10.20192195

**Authors:** Suhail Ganiny, Owais Nisar

## Abstract

India, the second-most populous country in the world, has been lately witnessing a daily surge in the COVID-19 infected cases. India is currently among the worst-hit nations worldwide, due to the COVID-19 pandemic, and ranks just behind Brazil and USA. In order to prevent the further worsening of the situation, predicting the future course of the pandemic is of utmost importance. In this paper, we model the past trajectory (March 01, 2020 - July 25, 2020) and make a month-long (July 26, 2020 - August 24, 2020) forecast of the future evolution of the COVID-19 pandemic in India using an autoregressive integrated moving average (ARIMA) model. According to our forecasting results, India is likely to have 3,800,989 cumulative infected cases, 1,634,142 cumulative active cases, 2,110,697 cumulative recoveries and 56,150 cumulative deaths by August 24, 2020, if the current trend of the pandemic continues to prevail. The implications of these forecasts are that in the upcoming month the infection rate of COVID-19 in India is going to escalate, while as the rate of recovery and the case-fatality rate is likely to reduce. In order to avert these possible scenarios, the administration and health-care personnel need to formulate and implement robust control measures, while the general public needs to be more responsible and strictly adhere to the established and newly formulated guidelines to slow down the spread of the pandemic and prevent it from transforming into a catastrophe.

## 1 Introduction

Since the first emergence of the Coronavirus Disease 2019 (COVID-19) in Wuhan, Hubei Province, China, in December, 2019 [1, 2, 3], the disease has proliferated globally and has affected 215 countries till date [4]. The causative agent of the disease has been identified to be novel Severe Acute Respiratory Syndrome Coronavirus 2 (SARS-CoV-2), that shares a 79.6% sequence match with its predecessor SARS-CoV [3]. SARS-CoV-2 is very likely to have a zoonotic origin, possibly from bats, as it has a similarity of 97% with SARS-like bat CoVs at the whole-genome level [2, 5]. It is however, quite probable that pangolins acted as the intermediate hosts prior to human transmission [6]. SARS-CoV-2 is the seventh pathogen belonging to the class of coronaviruses that tend to affect humans, the other six being HCoV-229E, HCoV-OC43, HCoV-NL63, HCoVHKU1, SARS-CoV and MERS-CoV [7].

The human-to-human transmission of COVID-19 predominantly occurs through respiratory droplets (5 *µm <* size *<* 10 *µm*) or aerosols (size *≤* 5*µm*), close interpersonal physical contact or by touching infected surfaces [8]. Very recently, Morawska and Cao [9] have also achnowledged air borne transmission of COVID-19. Once a person is infected, it is typically characterized by symptoms like fever, cough, fatigue, myalgia, chest pain, dyspnoea and sore throat [10]. The global outbreak and the severity of this contagious disease (transmissibility rate - *R*_0_ = 1.4-3.9 [11]) prompted the World Health Organization (WHO) to declare it as a Public Health Emergency of International Concern (PHEIC) on January 30, 2020, and then subsequently, to classify it as a pandemic on March 11, 2020 [12]. Moreover, the COVID-19 *R*_0_ is estimated to be as high as 5 for events like weddings, religious gatherings, conferences and in industrial settings [13], and thereby such events have, and continue to, accelerate the propagation of the disease. Owing to non-availability of a specific vaccine, the management of the disease requires the adoption of measures like social distancing, frequent hand washing and sanitizing, wearing face masks at individual level and extensive screening/testing, contact tracing, isolation and quarantine at the administrative level [14].

According to the current statistics (July 28, 2020, 18:48 GMT), COVID-19 has hitherto affected more than a 16 million people (16,779,951) globally, including greater than 10 million survivors (10,333,138), while more than 0.6 million people (660,318) have unfortunately succumbed to the disease [4]. The recovery rate and the case-fatality rate currently stand at 61.58% and 3.93%, respectively. The world is presently witnessing a daily surge of nearly 0.2 million newly infected cases and about 5,000 fatalities. The USA, Brazil, India, Russia and South Africa are being the five most worst affected nations at this moment with a case share of 27%, 15%, 9%, 5% and 3% of the total global cases, respec-tively. While as, the proportion of deaths in these countries is 23%, 14%, 6%, 2% and 1% of the total worldwide deaths, respectively. An overview of the relative spread of the pandemic among the ten heavily effected nations in the world is depicted in Fig. 1. The spread and fatality of this continuing pandemic differs stochastically from country to country as a multitude of factors like government response, economic status, testing rate, healthcare infrastructure, environmental conditions, demographics, faithful reporting, compliance with advised measures etc. contribute to it [15, 16]. The rapid outbreak of the pandemic has disrupted the normal life of the human inhabitants as we are being strictly adhering and adapting to measures like indoor confinement, nationwide lockdowns, social distancing, travel restrictions, administrative surveillance, limited outdoor activity owing to the closure of workplaces, educational institutes, restaurants, parks, gymnasiums etc. [17, 18].

**Figure 1:**
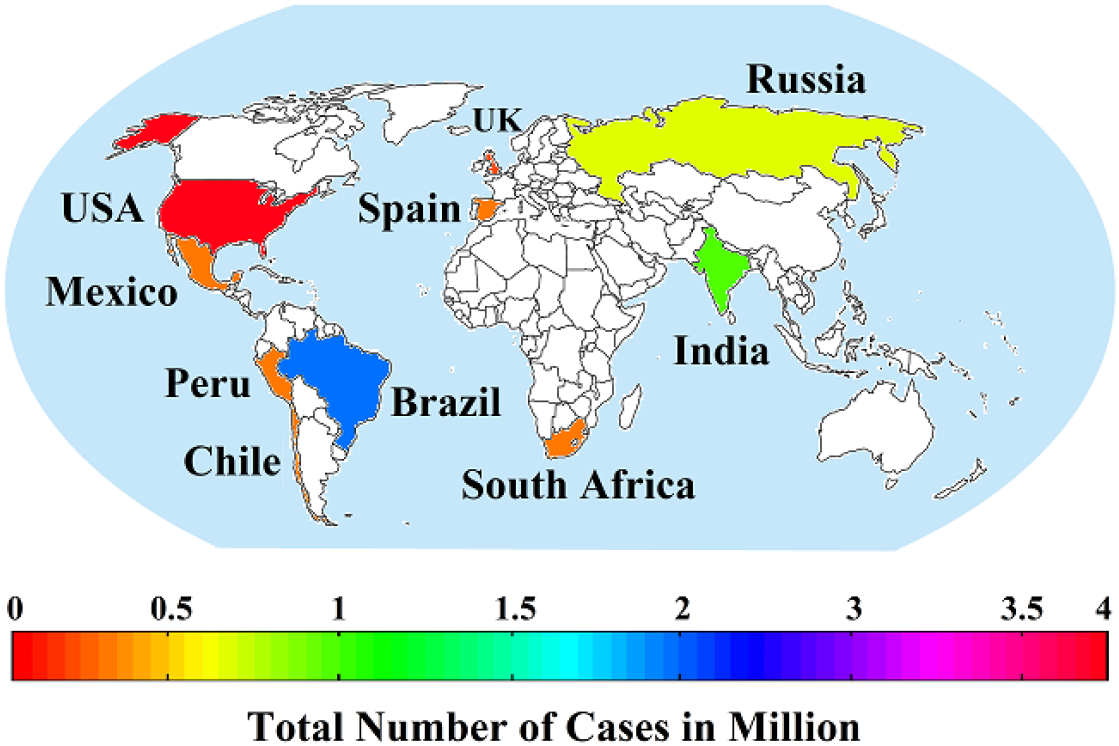
The relative spread of the COVID-19 pandemic in the top 10 worst affected countries as on July 28, 2020.

The first case of COVID-19 in India was reported on January 30, 2020, in Kerala (a coastal state in the southwestern part of India), when a student returnee from Wuhan, China, tested positive for the virus [19]. The second and the third cases were reported from the same state on February 2, 2020 and February 3, 2020, respectively, with both the patients having a Chinese travel history. The ongoing outbreak, however, started in March, 2020, when the indigenous cases started surfacing. India has ever since seen a continuing daily increase in the newly identified cases, the current daily count being nearly 50,000. Although, initially the spread of the disease was relatively slow, but gradually the transmission started picking up pace and as of now (July 28, 2020, 18:48 GMT), the total number of diagnosed cases has surged past 1.5 million (1,532,125). The diagnosed cases include about 1 million recoveries (988,768), more than 0.5 million active cases (509,133) and 34,224 deaths [4, 20]. With such high numbers, India is currently the worst affected Asian country and the third worst hit country worldwide by the pandemic and it accounts for nearly 38% of the identified cases in Asia and about 9% of the global cases. India currently has the highest infection rate globally as is evident by the fact that the cumulative infected cases have increased nearly by 20% since the last week and by 65% since the past one month only. India’s journey from the first inception of the virus till now is depicted in Fig. 2. India is emerging as the latest global hotspot of the pandemic owing to the high rise in the cases and the fact that most of the population of the country lives in densely packed cities. The recovery rate of 64.53% and the case-fatality rate of 2.23% in India are, however, promising and are better than the global values of 61.58% and 3.93%, respectively. It is pertinent to mention that the actual number of infected cases and the mystical low number of fatalities could in reality be much higher as India’s testing rates are one of the lowest (12,848 per million population [4]). Besides this, the relatively poor health-care infrastructure of the country, and the fact that a large proportion of the population dies in rural areas without any significant medical contemplation, renders their determination and reporting less likely.

**Figure 2:**
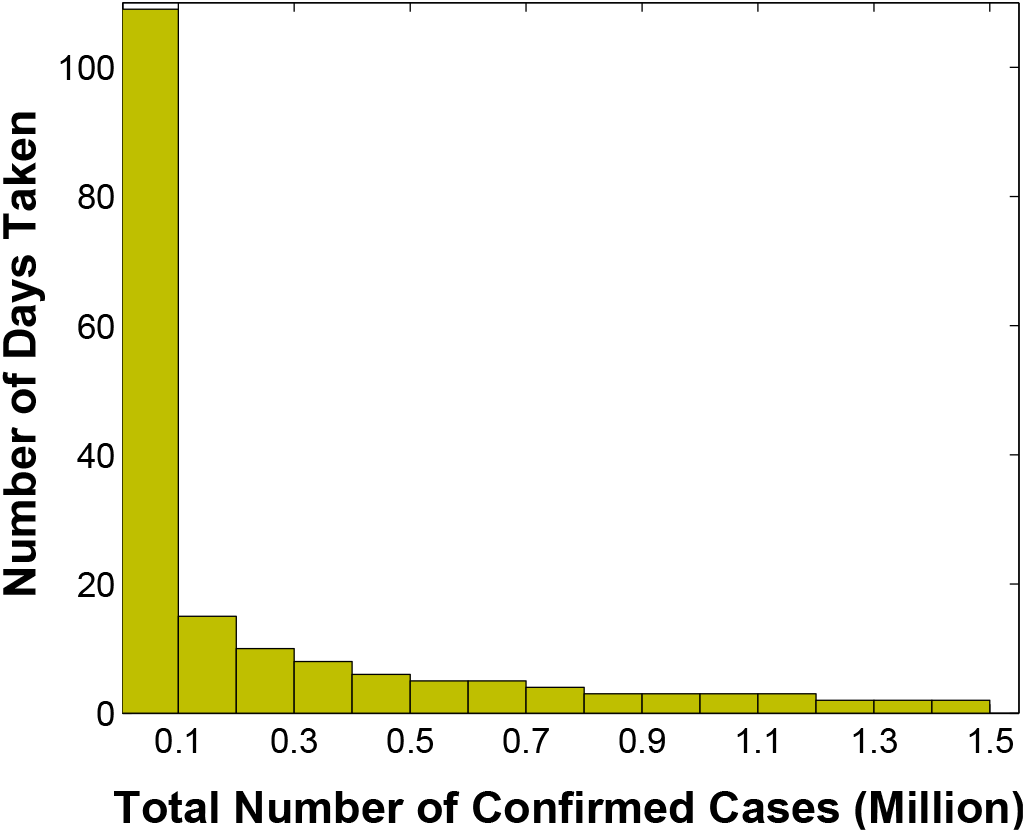
India’s Journey to 1.5 million cases – India took 109 days to reach 0.1 million, followed by 15 days to 0.2 million and just 2 days to reach from 1.4 million to 1.5 million.

COVID-19 has swept across all the 28 states and 8 union territories of India. The infected cases have particularly escalated in the sates of Maharashtra (391,440), Tamil Nadu (227,688), Andhra Pradesh (110,297), Karnataka (107,001), Uttar Pradesh (73,951), West Bengal (62,964), Gujarat (57,982), Telangana (57,142) and Bihar (43,591) [20]. These 10 worst affected states account for the almost 74% of the total diagnosed cases in India. The Indian Government has not yet declared the community transmission of COVID-19 in the country, however, the states of Assam, Kerala and West Bengal have announced the same [21]. In contadiction to this, many experts are of the opinion that the country is indeed in the community transmission phase of the pandemic as the sources of many diagnosed patients cannot be traced [22].

The Indian Government took several countermeasures initially to prevent the spread of the disease and to equip the hospitals and administration in order to be better prepared to deal with the pandemic. Beginning with the thermal screening of the passengers at airports, travel restrictions both domestically and internationally, closure of schools, workplaces, places of worship etc., encouraging people to practice social distancing and wearing mask and a one day voluntary nationwide shutdown [23, 24]. More stringent measures were enforced subsequently, starting with the imposition of a three week long nationwide lock-down on March 25, 2020, which was progressively extended quadruple times each with a duration of two weeks [25]. A brief timeline of the COVID-19 pandemic in India is shown in Fig. 3. Being the second largest populous country of the world (inhabited by 1.3 billion people), and the fact that the pandemic is yet to slow down its spread, it becomes extremely important to know how the trajectory of the disease is likely to evolve in the country. There are apprehensions that the pandemic could potentially lead to a high surge in number of infected people and the deceased and transform into a catastrophe, if the spread is not mitigated and effective control measures are not implemented. Thus the prediction of the ensuing course of the disease is very important as the outbreak further unfolds in the near future. Mathematical modeling of the pandemic is thus essential as the forecasts based on such models would prove immensely useful to policy makers, administration, health-care personnel etc. and help them to formulate various strategies and be in a state of preparedness to deal with eventualities that may be inevitable. Modeling and forecasting of the pandemic coupled with the strict following of the guidelines are extremely important until a dedicated vaccine is developed for the disease.

**Figure 3:**
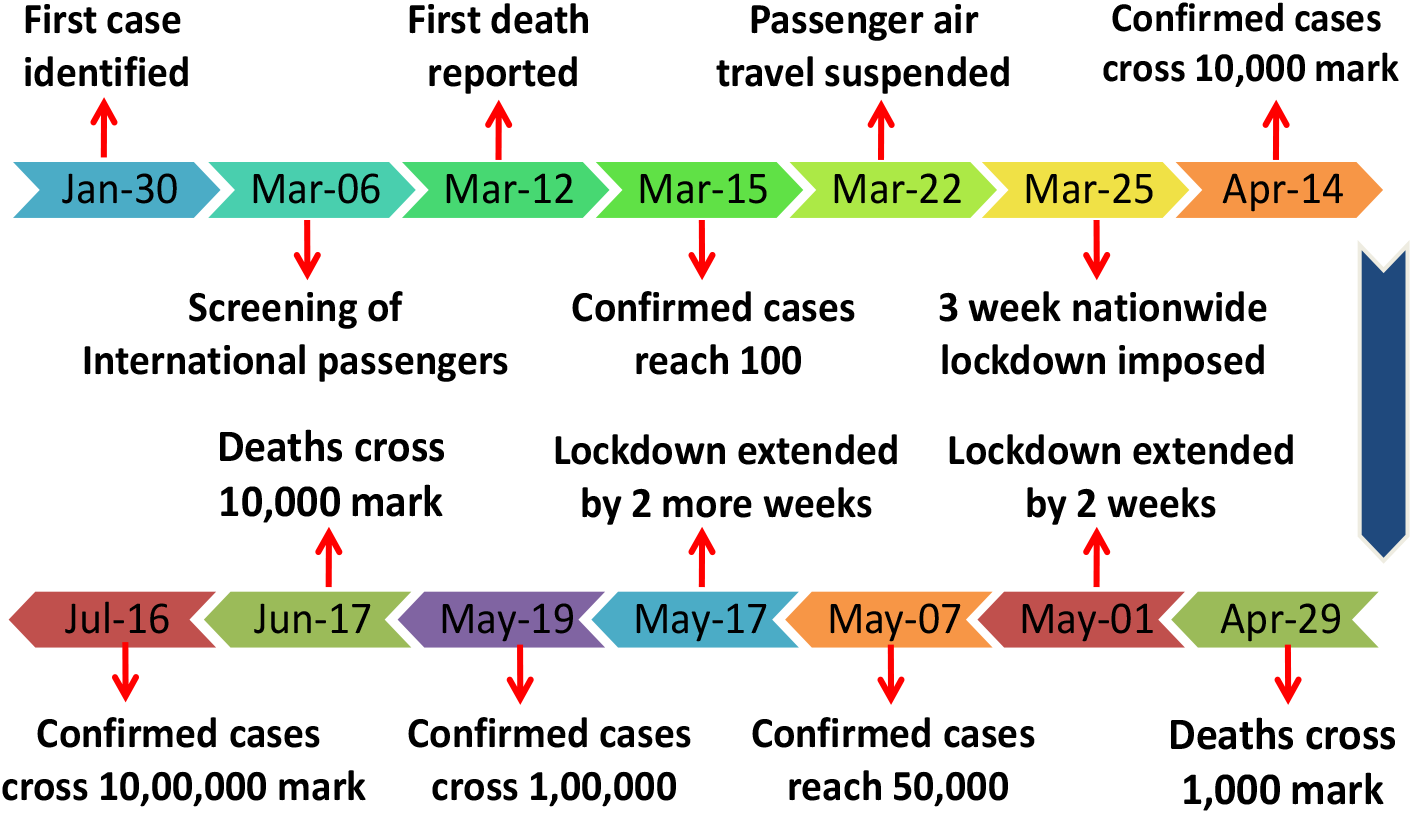
A brief timeline of the COVID-19 pandemic in India, depicting the milestones with respect to the administrative measures taken, and the pandemic statistics.

Several researchers have modeled the COVID-19 pandemic and forecasted its future evolution. For instance, among the first works in this regard Lin *et. al* [26] used an extension of the Susceptible-Exposed-Infectious-Removed (SEIR) compartmental model to account for the zoonotic origin of the COVID-19, individual response, governmental intervention and the emigration of the people in the early stages of the pandemic in Wuhan, China. A similar line of approach has been followed in [27] by Giordano *et. al*., their model SIDARTHE is also an extension of the SEIR model and includes other classes of people like diagnosed, ailing, threatened and healed. The SIDARTHE model has been used to model the COVID-19 spread in Italy. Anastassopoulou *et. al*. [28] estimated the transmission rate (*R*_0_), case-fatality rate and recovery rate based on an SIDR model for Hubei province, China, from January 11, 2020 - February 10, 2020 and also provided a three week long forecast of these epidemiological parameters. The transmissibility of the COVID-19 from super spreaders (an individual or a mass gathering) has been modeled in [29]. Hellewell *et. al*. [30] have modeled the effectivenss of isolation of infected patients and contact tracing on the COVID-19 spread. Their simulation results suggest that an extensive contact tracing and isolation of suspected cases is very likely to suppress the pandemic within few months, if the infections start spreading after the onset of symptoms. Eikenberry *et. al*. [31] have modeled the effectiveness of using face masks on the spread of the virus. Their results demonstrate that a broad use of face masks can prevent the widespread transmission of the disease and thereby reduce hospitalization of patients and fatalities. Rodriguez *et. al*. [32] have modeled the spread of the COVID-19 outbreak in Mexico using the Gompertz and Logistic models, and the machine learning approach based on the artificial neural network. Furthermore, the authors used the model inversions to extrapolate the unfolding of the disease for a duration of one week. In [33], the authors have used autoregressive integrated moving average (ARIMA) and nonlinear autoregressive artificial neural networks (NARANN) based approaches to analyze the prevalence of the pandemic in the African country of Egypt, using the data obtained from the Egyptian ministry of health (MoH). The findings of the study reveal that NARANN is statistically better in modeling the behavior of the pandemic for the Egyptian data. The authors in [34], have forecasted the number of cases, number of recoveries and the number of deaths for a one month period for Pakistan using ARIMA models and provided certain guide-lines in order to contain the spread of the virus. The modeling and provision of future projections in Nigeria, Saudi Arabia, Brazil, Canada, USA, Japan and Australia have been undertaken in [35, 36, 37, 38, 39, 40, 41].

Modeling and forecasting of the COVID-19 pandemic in India has been attempted by Sarkar *et. al*. [24], using *SARII_q_S_q_* model, which is a modified and a more general form of the SEIR model and takes into account the asymptomatic disease carriers (A), infected individuals who are isolated (*I_q_*) and susceptible individuals who are under quarantine (*S_q_*), besides taking the susceptible (S), recovered (R) and infected (I) persons into consideration. The authors have examined the reproduction rate (*R*_0_) of the spread and how it varies with administrative measures like lockdowns. They have also simulated the life-cycle of the disease and obtained its possible date of termination. However, owing to the limited data used in the analysis, the findings of the work are not consistent with the prevailing scenario. For instance, their simulation results were suggestive that the pandemic would terminate on July 26, 2020, however, as on this date, it is still far from being over, and in fact, as mentioned earlier, India is currently witnessing a daily increase of about 50,000 cases. A short term forecast (May 1, 2020 - May 22, 2020) of COVID-19 cases in India has been done in [42], based on an SIR model and a logistic growth model, with the latter been found to be more effective than the former. The three week projections of their analysis, as compared to the actual data, however, turned out to be significantly lower. The study also reveals that the lockdowns that were enforced in the country to suppress the spread of the disease did not had any statistically significant effect on the mitigation of the transmission. Tomar and Gupta [43], have used long short-term memory (LSTM) technique and the classical curve fitting for modeling and forecasting COVID-19 in India using a very limited data.

In this paper, we attempt to model the emergence of the COVID-19 pandemic in India, and to obtain a one month long forecast based on the developed model(s). We model and forecast: (1) Total number of diagnosed cases, (2) Total number of recoveries and (3) Total number of deaths, based on the publicly available data from March 1, 2020 to July 25, 2020. In addition to this, we also forecast the total number of active cases based on the forecasts obtained for the other three categories. Knowing that the pandemic in India has not shown any signs of slowing down yet, and is still monotonously increasing, we assume that the trend will prevail in the upcoming month. Our forecasts therefore represent the worst case numbers of the likely infected cases, recoveries and casualties that can be anticipated. The rest of the paper is organized as follows: In the next section, the rationale for modeling and prediction of COVID-19 is first presented, followed by the description of the data sources and finally the mathematical preliminaries of the modeling technique are detailed. In section 3, the results of the modeling and forecasting are provided and discussed briefly, finally the paper is concluded in section 4.

## 2 Modeling and Forecasting of COVID-19 in India

In this section, we mathematically model the progression of the COVID-19 pandemic in India from March 01, 2020 till July 25, 2020. We select a particular model within a certain class of models based on the statistical properties viz root-mean squared error, mean absolute error, coefficient of determination etc. and then attempt to predict the future course of the disease for a period of one month i.e., upto August 24, 2020. The modeling and forecasting of the COVID-19 pandemic is formulated as a typical univariate time-series problem using the autoregressive integrated moving average (ARIMA) technique, wherein, it is assumed that the current or future values of the diagnosed cases/recoveries/deaths are functions of their lagged (past) values. ARIMA models are typically used to model and forecast processes that yield a time-series as output, and have been used in varied areas ranging from transportaion forecasting [44], fuel energy demand forecasting [45], to Stock price prediction [46]. In order to develop the model, the available datasets are divided into two subsets: (1) Training data (90%), and (2) Validation data (10%). The training data is used to determine the unknown model parameters and the validation data is used to assess the forecasting capabilities of the developed models. A total of 29 models are obtained amongst which only one model is finally chosen that has better statistical characteristics than the others. The selected model is then used for forecasting month ahead projections from July 26, 2020 to August 24, 2020 for the total number of diagnosed cases, total number of recoveries and total number of deaths. Once these forecasts are obtained, the forecast for total number of active cases is obtained using:

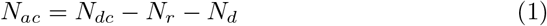

where,

*N_ac_* = Total number of active cases
*N_dc_* = Total number of diagnosed cases
*N_r_* = Total number of recoveries, and,
*N_d_* = Total number of deaths

### 2.1. Data Source

The data utilized in this study has been obtained from several reliable sources that include the the Government of India’s Ministry of Health and Family Welfare (MoHFW) [20], the website worldometer [4] and the website covidindia [47]. The data has been collected from March 1, 2020, till July 25, 2020, and pertains to the cumulative number of diagnosed (infected) cases, cumulative number of recovered patients, and cumulative number of the deceased patients. The cumulative number of active cases was determined using Eq. 1. The data was preprocessed using the MATLAB environment, and any anomalies, if present, were accordingly verified and corrected. The data is illustrated in Fig. 4..

**Figure 4:**
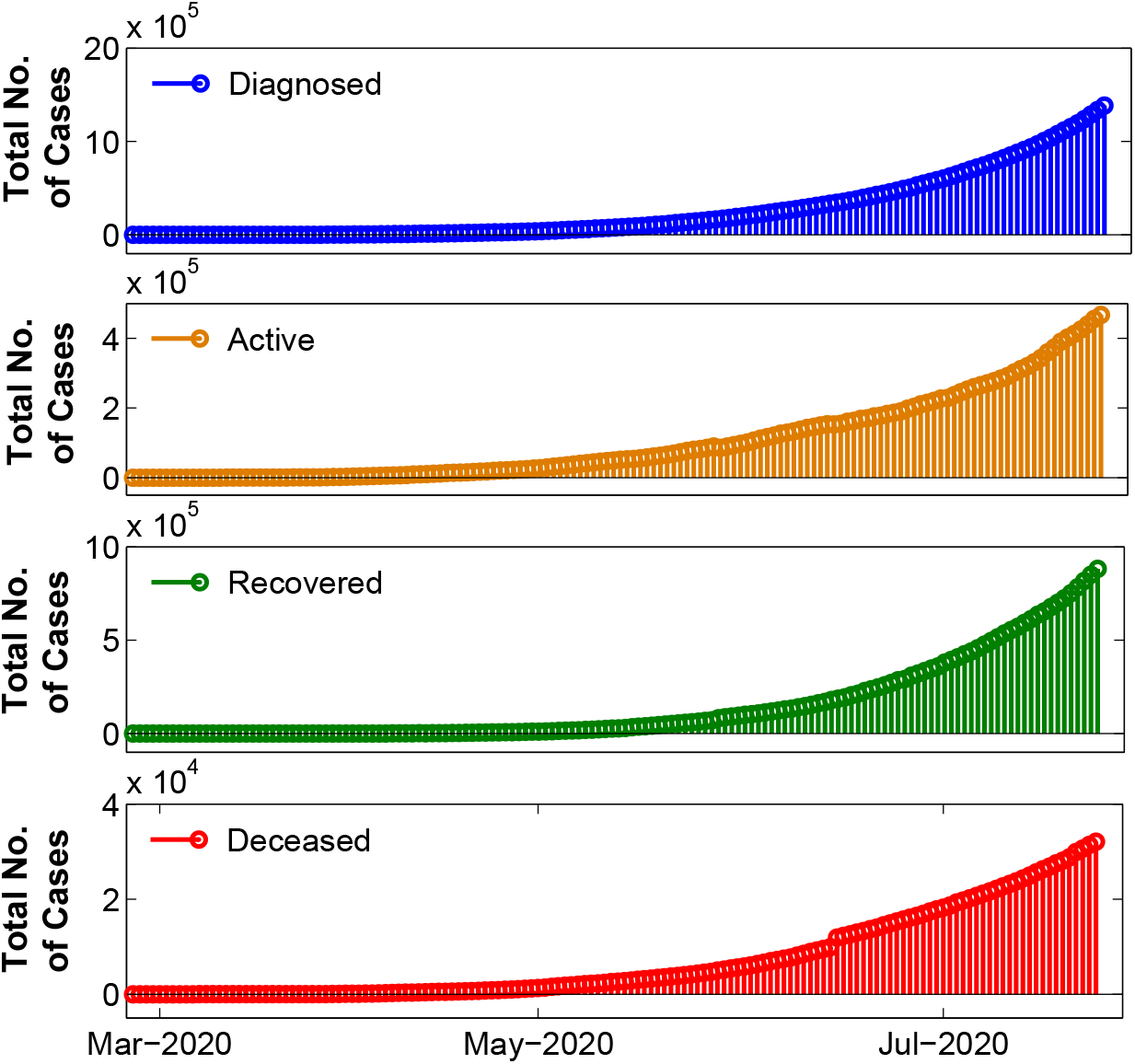
COVID-19 cumulative statistics in India, from March 01, 2020 to July 25, 2020. The data has been compiled from [4, 20, 47].

### 2.2. Mathematical Preliminaries

In order to make this paper self-contained, a brief introduction to time-series analysis and forecasting using ARIMA(p,d,q) model is essential, and the same is presented here. For an in-depth coverage of these topics, the interested readers can refer to the classical books [48, 49].

A time-series is a collection of data-points which are measured or observed after successive fixed time durations, for e.g, hourly, daily, weekly, monthly or annually [48]. Time-series are used to model and forecast various phenomenon ranging from epidemics/pandemics, weather, stock markets, transportation, etc. [34, 45, 46]. There are several models that can approximate their behavior and forecast their future evolution, for instance, exponential smoothing method, ARIMA(p,d,q), artificial neural network, logistic regression, etc [32, 42, 49]. ARIMA(p,d,q), however, remains by far, the most widely used.

The ARIMA(p,d,q) model comprises of the autoregressive part i.e., AR(p), and the moving average part i.e., MA(q), which have degrees *p* and *q*, respectively. The parameter *d* represents the order of differencing that is needed to stationarize the time-series. The optimum degrees, *p, d* and *q*, of the ARIMA(p,d,q) model can be determined using Akaike information criterion (AIC), corrected AIC, Bayesian Information Criterion (BIC) or from the autocorrelation function(ACF) plots and the partial autocorrelation function (PACF) plots [48, 49, 34]. Besides these three hyper-parameters, the ARIMA(p,d,q) models has certain unknown coefficients that are usually determined using the maximum likelihood estimation or the least square estimation techniques [48, 49].

A univariate discrete time-series is often represented as:

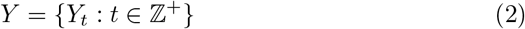

which can be written in expanded form as:

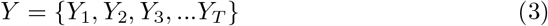

where, T is the total number of data-points in the time series.

In an ARMA(p,q) model, any general term of a nonseasonal and stationary time-series, 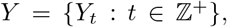, is assumed to be linearly dependent on the previous terms of the series, i.e.,

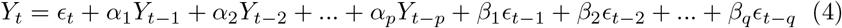

where, the *α′ s* represent the unknown coefficients in the AR(p) part of the model, and the *β′ s* are the unknown coefficients of the MA(q) part of the ARMA(p,q) model, respectively. The *∊′ s* denote the error terms, that are generally assumed to be normally distributed white noise signals with zero mean and a finite variance 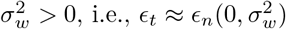 [48, 49].

In compact form, the ARMA(p,q) model assumes the form:

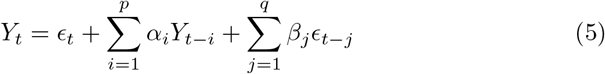

The ARMA(p,q) model is often specified in an alternate form, that requires the use of the backshift operator or lag operator, that is defined as:

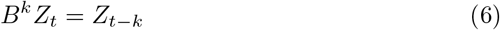

In terms of the backshift operator, Eq. 3 can be written as:

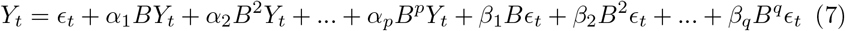

This can be put as:

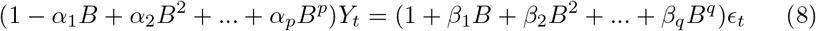

In compact form, this can be written as:

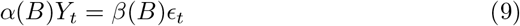

The ARMA(p,q) models approximate the time-series behavior, only if, the series is stationary, that is, when the statistical properties of the series are independent of the time interval in which they are observed. The following definitions [48, 49] of stationarity are often used in connection with time-series:

1. A time-series 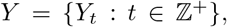 is said to be strictly stationary, if the statistical properties of (*Y_t1_*, *Y_t2_*, *Y_t3_*…*Y_tn_*) remain invariant under a time shift operation. In other words, if the statistical properties of (*Y_t1_*, *Y_t2_*, *Y_t3_*…*Y_tn_*) are exactly same as those of (*Y_t1+τ_*, *Y_t2+τ_*, *Y_t3+τ_*…*Y_tn+τ_*) *∀ τ* This is however a very strong condition and difficult to verify analytically for a time-series.
2. A time-series 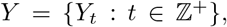 is said to be weakly stationary, if both the mean of the series, *Y*, and the covariance of the series terms, *Y_t_* and *Y_t−m_*, exhibit invariance with respect to time. More specifically, a time-series is weakly stationary if:

i. The mean of the time-series 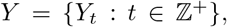 is a constant, i.e, *E*(*Y*) = *µ* (a constant), and
ii. The covariance between the terms *Y_t_* and *Y_t−m_* with a window length of *m* is only a function of *m*, i.e, *Cov*(*Y_t_, Y_t−m_*) = *η*(*m*).

In practice, the weak stationarity of a time-series is often visually assessed by the plot of time-series data. If the plot appears to fluctuate about a certain mean value, then the series is stationary. Similarly, if the autocorrelation functions of the time-series exhibit a decaying trend then the series is stationary. A non-stationary time-series cannot be modeled by ARMA(p,q) model directly, unless the series is first stationarized. A non-stationary series is often made stationary by using differencing, in which a transformation of the following form is applied on the series:

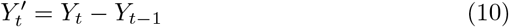

The order of differencing (*d*) needed to stationarize a time-series, varies as per the nature of the series. The ARMA(p,q) model coupled with a prior differencing is collectively known as ARIMA(p,d,q) model. The parameter estimation (*α*’s and *β*’s) and the forecasts based on the developed models are usually performed using dedicated programming routines of the commercially available software packages like in MATLAB, Mathematica, Python, R and Stata. We have, in fact, used MATLAB for the purpose here.

### 2.3. Application to Indian COVID-19 data

A simple line plot of the total number of diagnosed cases is shown in Fig. 5, along with the first order differenced, and the second order differenced time-series of the same data. The data is normalized to aid in the visualization of the trends in the series, as their ranges are different. The monotonously increasing trend of the actual data and the first order differenced data implies the nonstationarity of the time-series. The second order differenced series, is however, stationary as its fluctuates about a mean value. Thus, to model this time-series using ARMA(p,q), and to obtain forecasts based on this, a prior differencing of atleast a second order is necessary. We have thereby chosen two values of *d* = 2 and 3, to model this time-series here.

**Figure 5:**
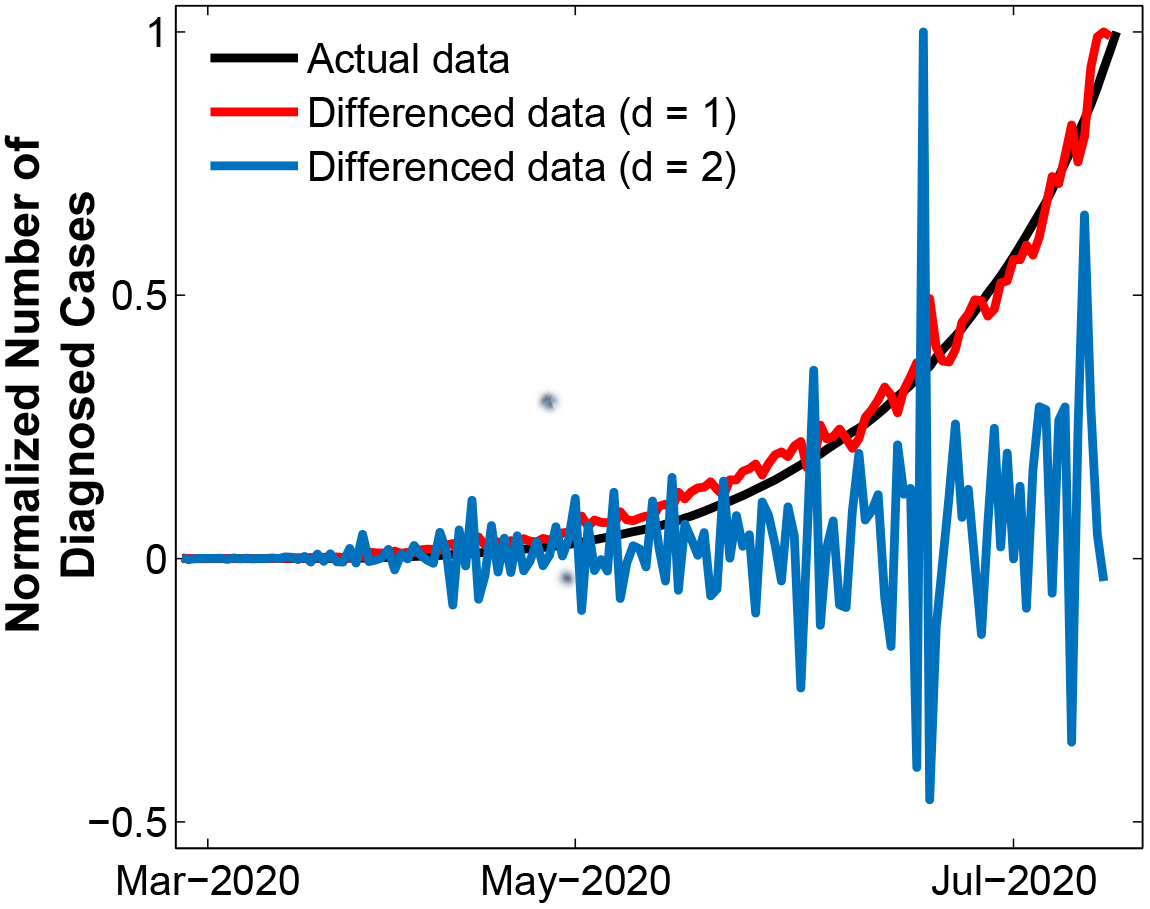
Normalized plots of the cumulative number of diagnosed cases, first order differenced data and the second order differenced data. The original series and the first order differenced series is non-stationary, while the second order differenced series is stationary.

In order to determine the likely values of the hyperparameters *p* and *q*, we resort to the ACF and PACF plots of the time-series, corresponding to the total number of diagnosed cases. The ACF and PACF plots are shown in Fig. 6. Since the ACF and PACF plots, neither show a gradual damping, nor a cutoff at a single value of lag. We use several arbitrary values for the parameter, *p* = 2, 5, 7, 8, 9, 14 and 15, and, the parameter, *q* = 2, 3, 5 and 7. An iterative procedure is thereby used for various combinations of *p, d* and *q* to determine the model that exhibits better statistical properties and has superior forecasting capabilities. The statistical metrics of root-mean-square error (RMSE), mean absolute error (MAE), mean absolute percentage error (MAPE) and the coefficient of determination (*R*^2^) are used for the model validation. The results of the iterations for the validation data set are depicted in Table 1. From this table, it can be clearly observed that the ARIMA(7,2,2) model, has the minimum values of the error metrics, and maximum value of the coefficient of determination, and therefore is the optimum model. We assume that for the other datasets similar inferences are true. It is important to mention that the missing combinations of the *p, d* and *q* values turned out to unstable and hence it was not possible to establish their statistical parameters.

**Figure 6:**
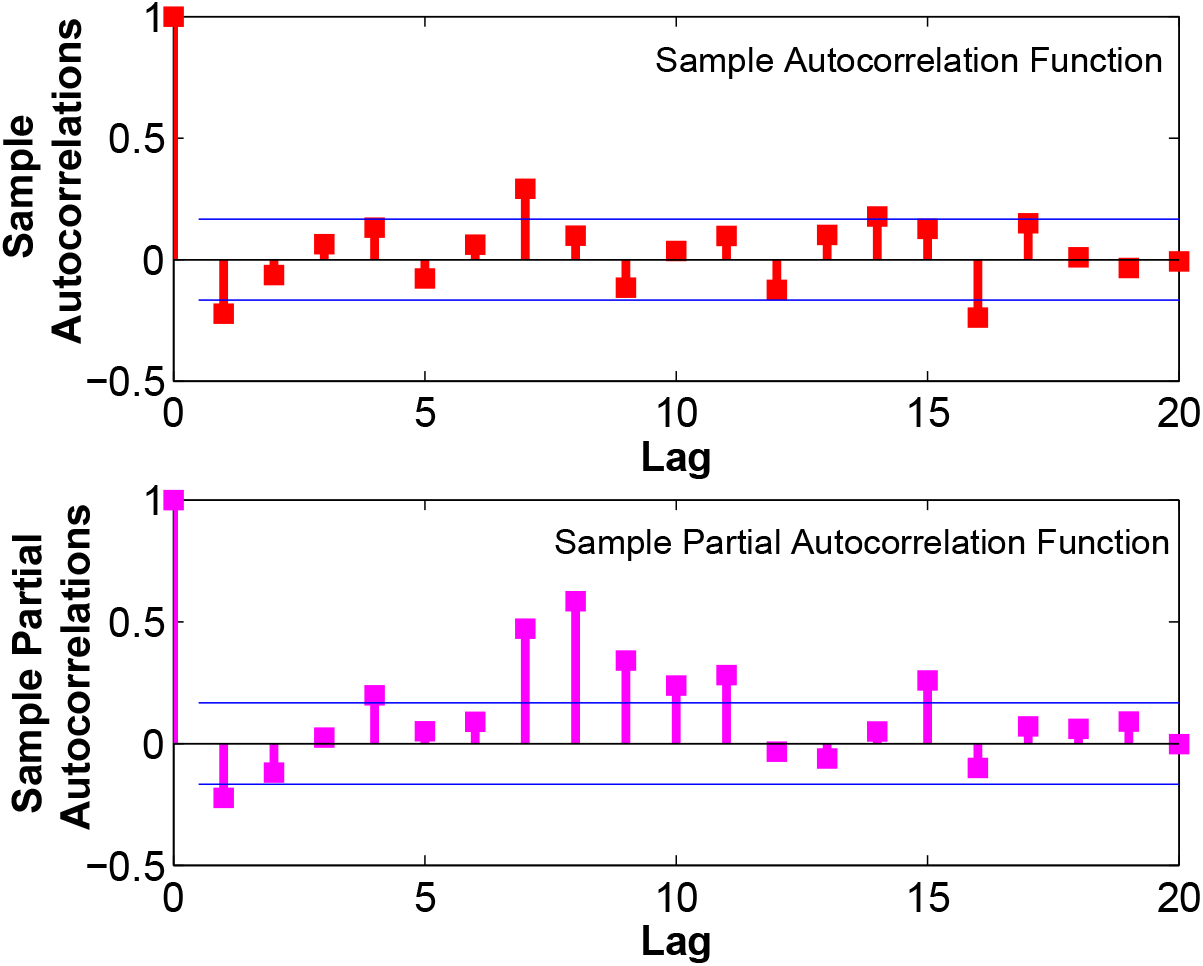
The autocorrelation function (ACF) plot, and the partial autocorrelation function (PACF) plot of the second order differenced series for the total number of diagnosed cases.

**Table 1:** Statistical metrics of the various ARIMA(p,d,q) models

| S.No. | ARIMA(p,d,q) model | RMSE | MAE | MAPE | $R^2$ |
| --- | --- | --- | --- | --- | --- |
| 01 | ARIMA(2,2,2) | 597.7 | 395.1 | 0.428 | 0.99997 |
| 02 | ARIMA(2,2,3) | 551.4 | 384.98 | 0.36986 | 0.99997 |
| 03 | ARIMA(2,2,5) | 542.68 | 368.57 | 0.29912 | 0.99998 |
| 04 | ARIMA(2,2,7) | 552.18 | 402.06 | 0.65024 | 0.99997 |
| 05 | ARIMA(5,2,2) | 534.29 | 390.61 | 0.59157 | 0.99998 |
| 06 | ARIMA(5,2,3) | 538.31 | 388.46 | 0.49523 | 0.99998 |
| 07 | ARIMA(7,2,2) | 457.61 | 330.79 | 0.2471 | 0.99998 |
| 08 | ARIMA(7,2,3) | 481.5 | 341.27 | 0.34807 | 0.99998 |
| 09 | ARIMA(7,2,5) | 473.14 | 345.13 | 0.3912 | 0.99998 |
| 10 | ARIMA(8,2,2) | 510.71 | 358.26 | 0.65241 | 0.99998 |
| 11 | ARIMA(8,2,3) | 481.48 | 341.38 | 0.35068 | 0.99998 |
| 12 | ARIMA(8,2,5) | 480.48 | 342.45 | 0.45999 | 0.99998 |
| 13 | ARIMA(9,2,2) | 507.76 | 355.99 | 0.67046 | 0.99998 |
| 14 | ARIMA(9,2,3) | 481.33 | 342.28 | 0.34025 | 0.99998 |
| 15 | ARIMA(9,2,5) | 472.24 | 340.38 | 0.45992 | 0.99998 |
| 16 | ARIMA(14,2,2) | 480.42 | 341.09 | 0.67289 | 0.99998 |
| 17 | ARIMA(15,2,2) | 514.69 | 357.46 | 0.62912 | 0.99998 |
| 18 | ARIMA(2,3,2) | 558.4 | 394.53 | 0.44165 | 0.99997 |
| 19 | ARIMA(2,3,5) | 545.43 | 383.91 | 0.37512 | 0.99997 |
| 20 | ARIMA(2,3,7) | 514.46 | 397.19 | 0.47946 | 0.99998 |
| 21 | ARIMA(5,3,2) | 523.06 | 374.22 | 0.3845 | 0.99998 |
| 22 | ARIMA(5,3,5) | 544.49 | 354.65 | 0.26738 | 0.99997 |
| 23 | ARIMA(5,3,7) | 472.74 | 333.65 | 0.39325 | 0.99998 |
| 24 | ARIMA(7,3,2) | 511.03 | 358.35 | 0.46553 | 0.99998 |
| 25 | ARIMA(7,3,5) | 472.91 | 344.74 | 0.35958 | 0.99998 |
| 26 | ARIMA(8,3,2) | 507.77 | 359.32 | 0.4316 | 0.99998 |
| 27 | ARIMA(8,3,5) | 472.53 | 343.8 | 0.3787 | 0.99998 |
| 28 | ARIMA(14,3,2) | 463.39 | 340.99 | 0.54049 | 0.99998 |
| 29 | ARIMA(15,3,2) | 468.69 | 342.67 | 0.39887 | 0.99998 |

## 3. Results and Discussions

The modeling and predicting capabilities of the ARIMA(7,2,2) models for the total number of diagnosed cases, total number of recoveries and the total number of deaths are illustrated in Figs 7, 8 and 9. It has to be recalled, that we had partitioned the available data from March 01, 2020 to July 25, 2020, into two groups: the first, for training purposes (90%), i.e., for model development, and the second, for validation purposes (10%), i.e., for assessing the predicting capabilities of the developed models. From Figs, it is clear that ARIMA(7,2,2) models have very good fitting (with respect to training data) and predicting (with respect to validation data) capabilities for all the three categories of datasets. The values predicted by the ARIMA(7,2,2) models are quite close to the actual values. The small deviations between the predicted values and the actual values have been quantified using the statistical metrics of RMSE, MAE, MAPE and *R*^2^, and are provided in Table 1, for the total number of diagnosed cases.

**Figure 7:**
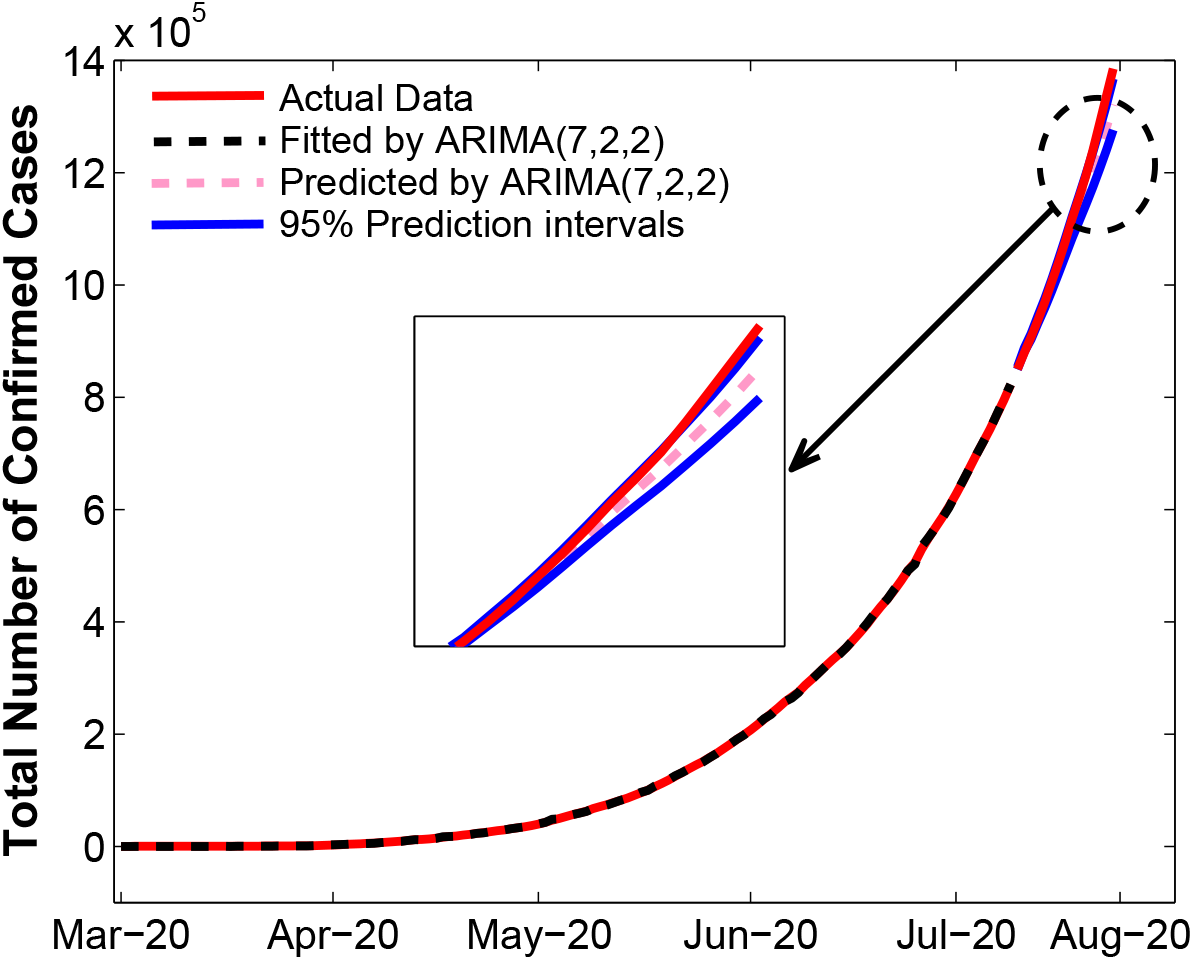
A comparison of the actual data pertaining to the cumulative number of diagnosed cases, and the data fitted by and predicted by the ARIMA(p,d,q) model.

**Figure 8:**
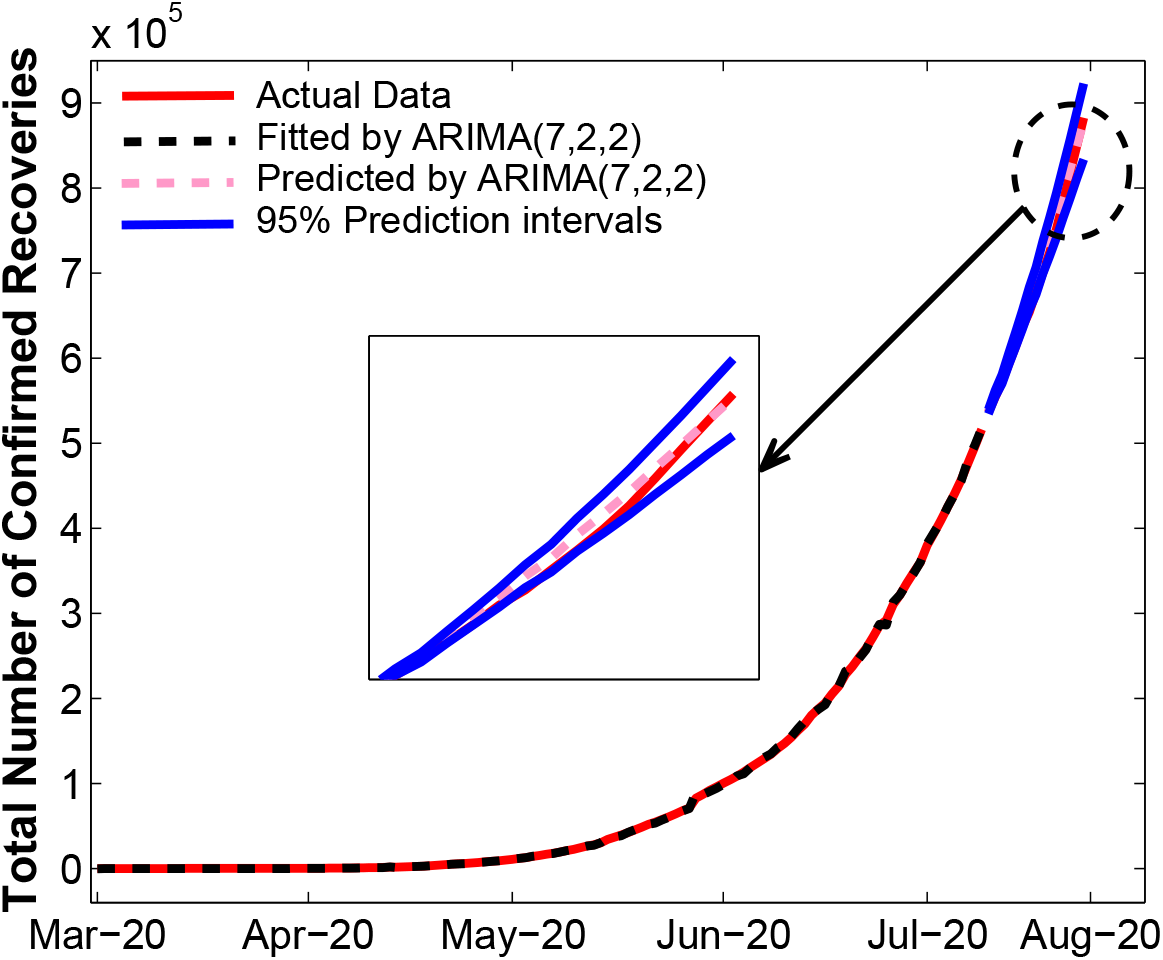
A comparison of the actual data pertaining to the cumulative number of recoveries, and the data fitted by and predicted by the ARIMA(p,d,q) model.

**Figure 9:**
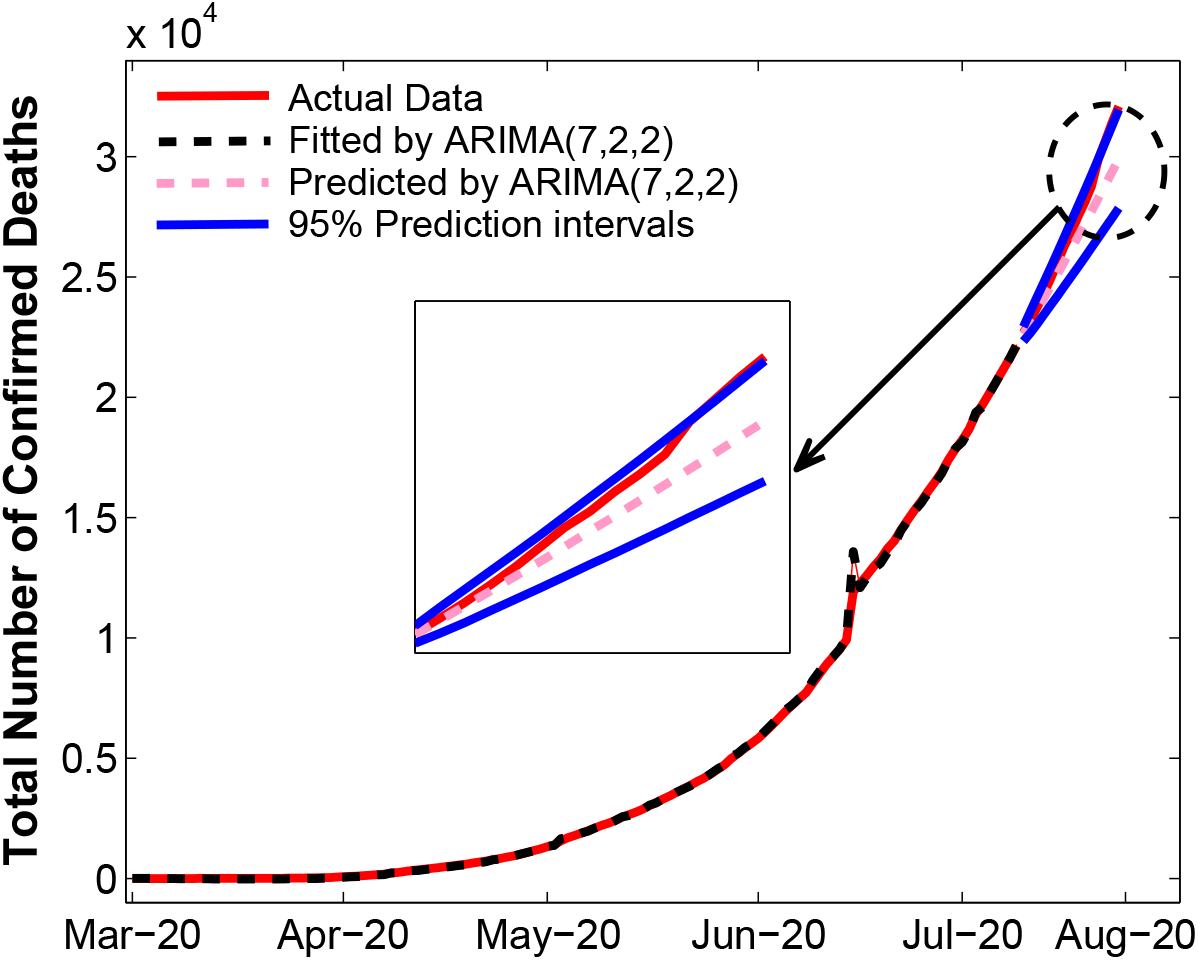
A comparison of the actual data pertaining to the cumulative number of deaths, and the data fitted by and predicted by the ARIMA(p,d,q) model.

The one month ahead forecasts using the ARIMA(7,2,2) models, for the total number of diagnosed cases, total number of recoveries and the total number of deaths are shown in Figs 10, 11 and 12, along with their 95% confidence intervals. The absolute values are provided in Table 2, along with the forecast for the total number of active cases. Our forecasts predict that by August 24, 2020, the expected number of cumulative diagnosed cases would increase nearly three-fold from now, and surge to 3,800,989, the number of recoveries would reach 2,110,697 and the cumulative number of deaths would mount to 56,150. India is likely to cross the two million diagnosed cases mark on August 5, 2020, and the three million cases on August 17, 2020. The cumulative recoveries are expected to breach the two million mark on August 22, 2020, and the cumulative deaths could hit the fifty thousand mark on August 17, 2020. The daily increment in cumulative diagnosed would be nearly 110,182, for recoveries it will be 45,914, and for deaths the value would be 874. These forecasts would also correspond to a recovery rate of 55.53% and case-fatality rate of 1.47%. In comparison with the current values of 64.53% and 2.23%, both the recovery rate and the case-fatality rate would be lower. While, a lower case-fatality rate is desirable, however, a lower recovery rate would be a matter of concern. With regard to the rate of COVID-19 spread in the upcoming month, it is found that the cumulative diagnosed cases would escalate by 63.54%, which is lower than the current value of 65% and therefore would be promising.

**Figure 10:**
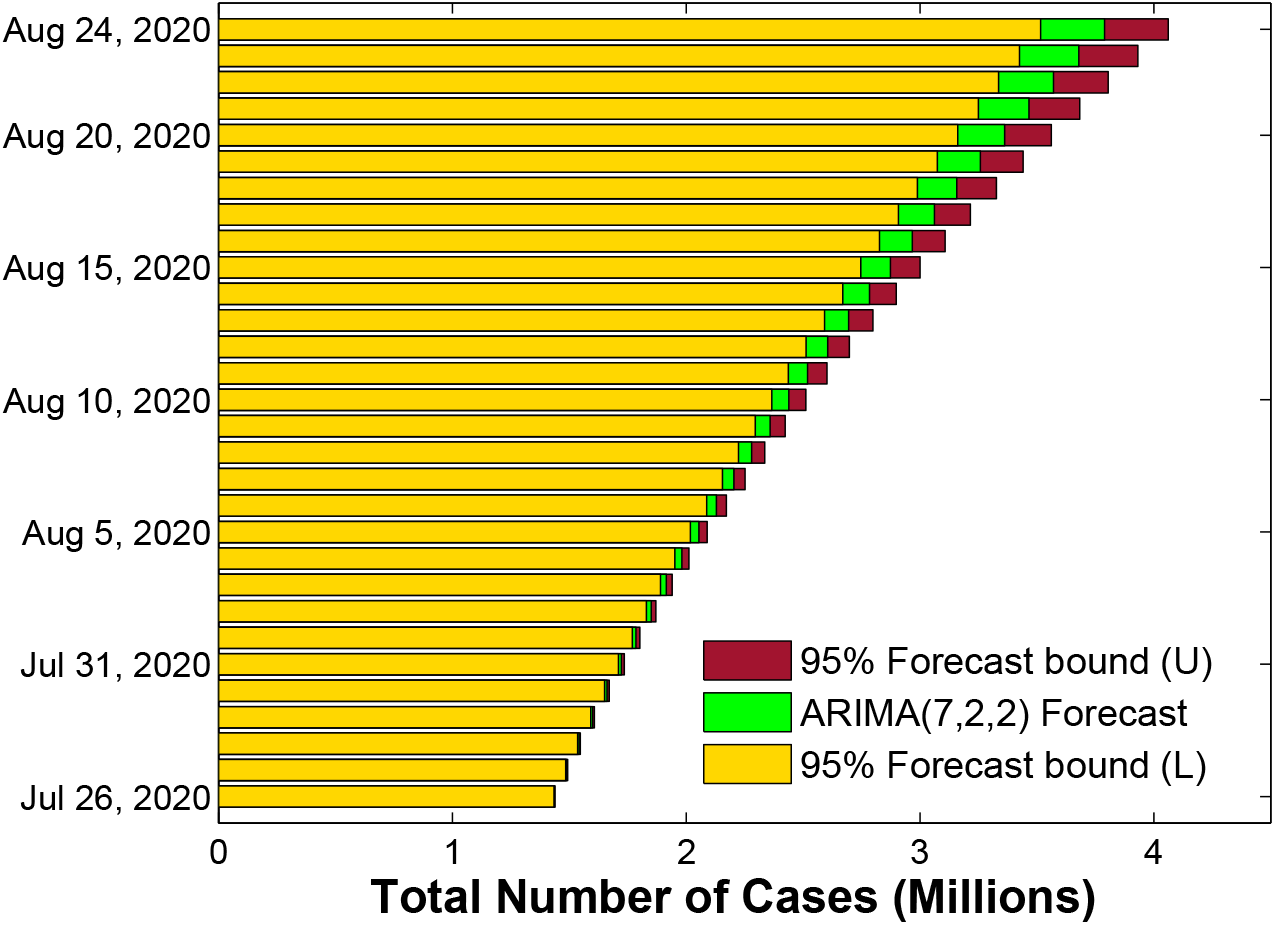
A one month forecast (July 26, 2020 - August 24, 2020), along with the 95% confidence limits, of the cumulative number of diagnosed cases using ARIMA(7,2,2) model.

**Figure 11:**
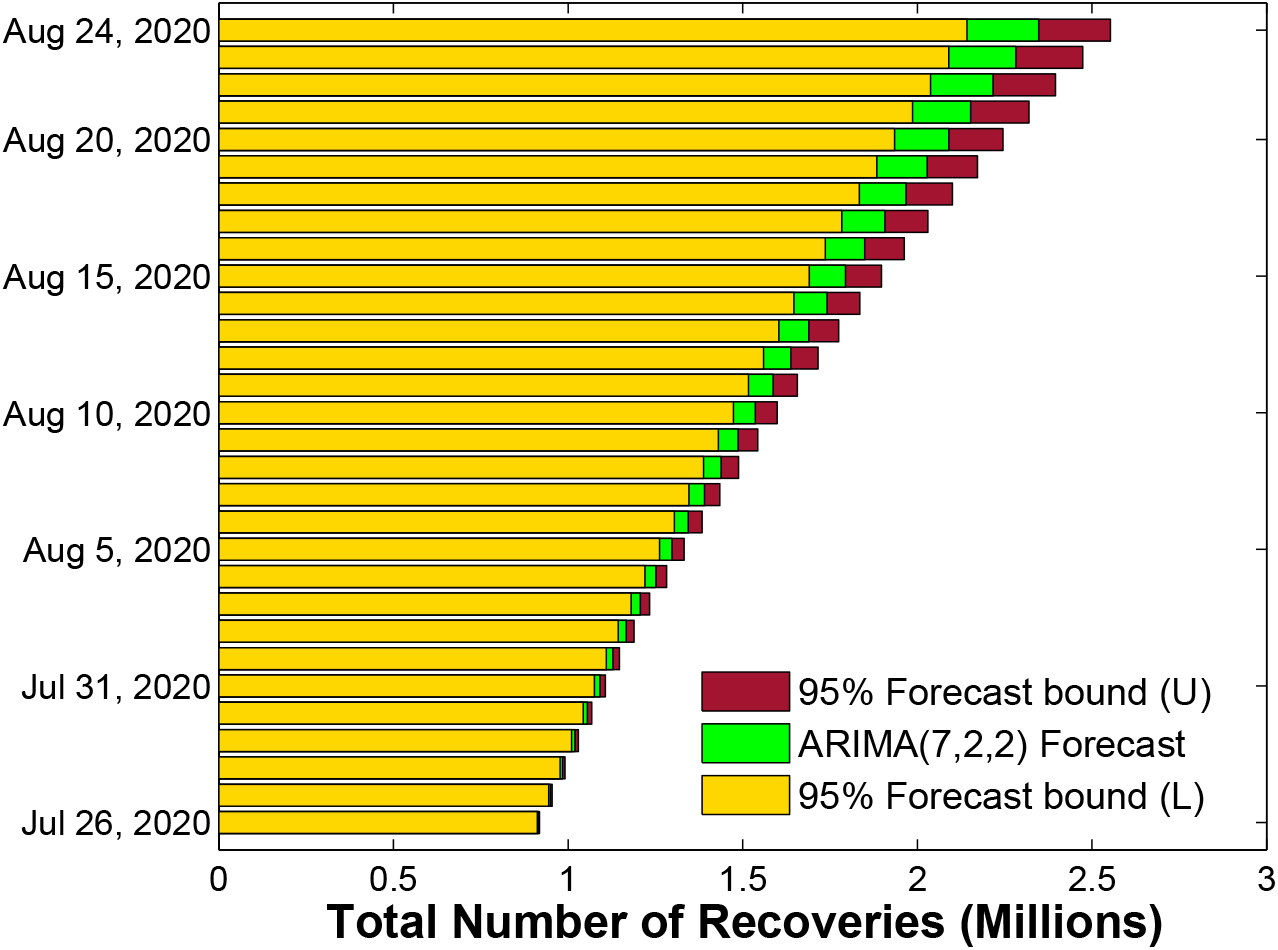
A one month forecast (July 26, 2020 - August 24, 2020), along with the 95% confidence limits, of the cumulative number of recoveries using ARIMA(7,2,2) model.

**Figure 12:**
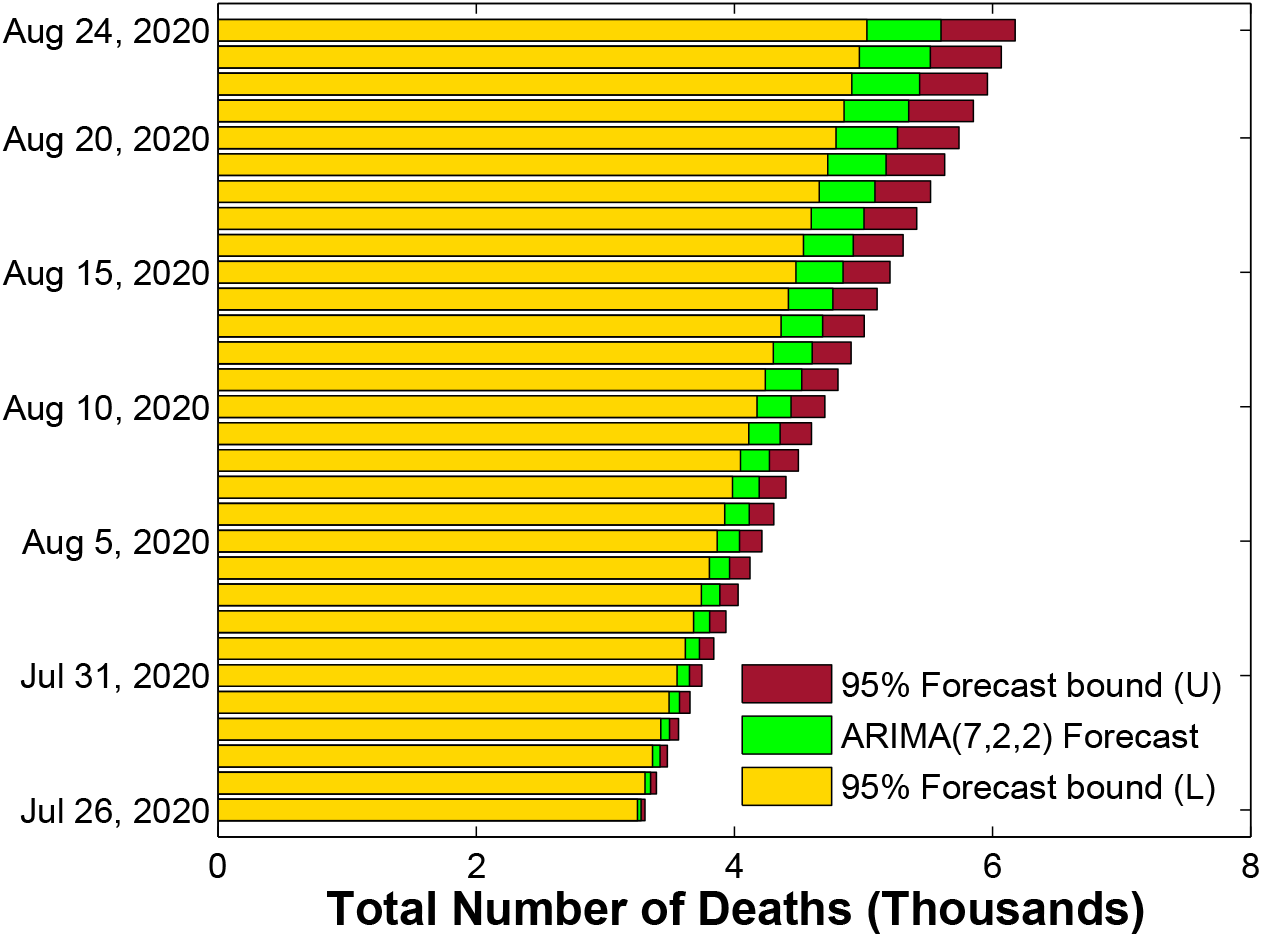
A one month forecast (July 26, 2020 – August 24, 2020), along with the 95% confidence limits, of the cumulative number of deaths using ARIMA(7,2,2) model.

**Table 2:** One month ahead forecast (July 26, 2020 – August 24, 2020) of total number of diagnosed cases, total number of recoveries, total number of deaths and total number of active cases in India. The values within the parantheses are the upper and lower 95% confidence limits.

| S.No. | Total no. of<br>diagnosed cases | Total no. of<br>recoveries | Total no. of<br>deaths | Total no. of<br>active cases |
| --- | --- | --- | --- | --- |
| 1 | 1436970 (1439742,1434197) | 916949 (920450,913448) | 32809 (33107,32510) | 487212 |
| 2 | 1488550 (1493153,1483946) | 952696 (958451,946942) | 33569 (34019,33118) | 502285 |
| 3 | 1542049 (1548455,1535643) | 989676 (998366,980987) | 34316 (34895,33737) | 518057 |
| 4 | 1599989 (1608489,1591490) | 1027654 (1039853,1015454) | 35076 (35776,34375) | 537259 |
| 5 | 1661463 (1672585,1650341) | 1065996 (1082302,1049691) | 35823 (36642,35004) | 559644 |
| 6 | 1723752 (1737844,1709660) | 1103893 (1125261,1082524) | 36573 (37524,35623) | 583286 |
| 7 | 1786352 (1804051,1768652) | 1141193 (1167754,1114633) | 37331 (38417,36244) | 607828 |
| 8 | 1850560 (1873047,1828074) | 1179167 (1211860,1146475) | 38093 (39321,36865) | 633300 |
| 9 | 1916205 (1944187,1888223) | 1218547 (1257883,1179212) | 38862 (40234,37491) | 658796 |
| 10 | 1984113 (2017926,1950300) | 1259177 (1305608,1212746) | 39635 (41152,38118) | 685301 |
| 11 | 2055690 (2095741,2015638) | 1300182 (1354122,1246242) | 40413 (42078,38747) | 715095 |
| 12 | 2130546 (2177436,2083657) | 1340729 (1402452,1279006) | 41195 (43013,39377) | 748622 |
| 13 | 2206770 (2261133,2152406) | 1380691 (1450816,1310567) | 41982 (43955,40009) | 784097 |
| 14 | 2283610 (2346262,2220959) | 1420456 (1499368,1341544) | 42775 (44907,40643) | 820379 |
| 15 | 2361733 (2433697,2289770) | 1460953 (1549155,1372752) | 43572 (45866,41279) | 857208 |
| 16 | 2441719 (2523874,2359564) | 1502730 (1600620,1404841) | 44375 (46834,41917) | 894614 |
| 17 | 2524282 (2617255,2431308) | 1545514 (1653395,1437632) | 45183 (47810,42556) | 933585 |
| 18 | 2610185 (2714570,2505799) | 1588414 (1706629,1470200) | 45996 (48793,43198) | 975775 |
| 19 | 2699103 (2815615,2582592) | 1630626 (1759429,1501824) | 46814 (49786,43842) | 1021663 |
| 20 | 2789688 (2919145,2660231) | 1672212 (1812007,1532417) | 47637 (50786,44488) | 1069839 |
| 21 | 2881174 (3024520,2737827) | 1713897 (1865060,1562735) | 48465 (51794,45136) | 1118812 |
| 22 | 2973945 (3132225,2815665) | 1756556 (1919453,1593660) | 49299 (52811,45786) | 1168090 |
| 23 | 3068765 (3242950,2894580) | 1800472 (1975416,1625527) | 50137 (53836,46439) | 1218156 |
| 24 | 3166347 (3357248,2975445) | 1845102 (2032324,1657881) | 50981 (54868,47094) | 1270264 |
| 25 | 3267126 (3475504,3058749) | 1889550 (2089311,1689790) | 51830 (55909,47750) | 1325746 |
| 26 | 3370749 (3597440,3144058) | 1933237 (2145792,1720683) | 52684 (56957,48410) | 1384828 |
| 27 | 3476193 (3722148,3230238) | 1976444 (2202116,1750773) | 53543 (58014,49071) | 1446206 |
| 28 | 3582794 (3849065,3316523) | 2020040 (2259157,1780923) | 54407 (59078,49735) | 1508347 |
| 29 | 3690807 (3978498,3403115) | 2064783 (2317638,1811928) | 55276 (60151,50401) | 1570748 |
| 30 | 3800989 (4111155,3490823) | 2110697 (2377543,1843852) | 56150 (61231,51070) | 1634142 |

The forecasts of this study can be helpful to the authorities to put effective control efforts in place and to take timely actions to contain the spread of the pandemic. Furthermore, the authorities can equip themselves with sufficient number of hospital beds, ventilators etc. and accordingly be well prepared to deal with the overwhelming of the hospitals. Although, the forecasts reported here, are based on the actual pandemic data, however, it is to be noted that these forecasts are subjected to many influencing factors and thereby the actual numbers could be different than the ones reported here. The most important factors would be the availability of vaccine, testing rates, adherence to measures like social distancing, hand-washing, sanitizing and wearing of face masks. Besides these, certain other factors that include, the phased relaxations offered by the local bodies and Government, economic conditions of individuals and the country as a whole and mass migrations of the people would also affect the numbers reported here.

## 4. Conclusion

In this paper, the authors modeled the past behavior and forecasted the possible future outcomes of the COVID-19 pandemic in India. The forecasting results reveal that the pandemic is likely to spread at a much faster rate while the recoveries are going to slow down and the fatality ratio is likely to reduce. The forecasted results are worrying and suggest that unless new control measures are devised/implemented and established guidelines are strictly followed the pandemic has the potential to turn devastating.

## Data Availability

The data used in the manuscript is available in public domain and the sources have been appropriately cited in the manuscript.

